# Radiological Evaluation of the Natural History of PIK3CA-Related Overgrowth Spectrum (PROS)

**DOI:** 10.64898/2026.03.09.26347786

**Authors:** Antoine Fraissenon, Gabriel Morin, Natalie Boddaert, Laureline Berteloot, Laurent Guibaud, Guillaume Canaud

**Affiliations:** INSERM U1151, Institut Necker-Enfants Malades, Paris, France; Centre de Référence des Anomalies Vasculaires Superficielles, Imagerie Pédiatrique, hôpital Femme-Mère-Enfant, HCL, Bron, France; CREATIS UMR 5220, Villeurbanne, France; Service de Radiologie Mère-Enfant, hôpital Nord, Saint Etienne, France; Université Paris Cité, Paris, France; Unité de Médecine Translationnelle et Thérapies Ciblées, Hôpital Necker-Enfants Malades, AP-HP, Paris, France; Centre d’Investigation Clinique – Unité de Recherche Clinique Mère-Enfant Necker-Cochin, Hôpital Necker-Enfants Malades, AP-HP, Paris, France; Service de Radiologie Pédiatrique, hôpital Necker-Enfants Malades, AP-HP, Paris, France

**Author notes:** Corresponding author: Prof. Guillaume Canaud, Université Paris Cité, Unité de médecine translationnelle et thérapies ciblées, Hôpital Necker-Enfants, Malades, AP-HP, Paris, France. Institut National de la Santé et de la Recherche Médicale U1151 149 rue de Sèvres, 75015, Paris, France. Contributed equally to the work (Antoine Fraissenon, Gabriel Morin). Contributed equally to the work (Laurent Guibaud, Guillaume Canaud).

## Abstract

*PIK3CA*-related overgrowth spectrum (PROS) comprises a heterogeneous group of disorders caused by postzygotic activating variants in *PIK3CA*, leading to mosaic activation of the PI3K pathway. PROS natural history is highly variable across patients and depends on the timing and distribution of the somatic mutation. To better characterize this natural history, we analyzed a cohort of patients with PROS.

This multicenter study was conducted at Hôpital Femme Mère Enfant (Lyon, France) and Hôpital Necker-Enfants Malades (Paris, France). We included pediatric and adult patients with PROS who had a documented *PIK3CA* variant and at least two MRI examinations performed at least 2 months apart. Patients undergoing interventional surgical or radiological procedures, or receiving targeted therapies were excluded. In all patients, target lesions were identified on baseline MRI scans, and assessed on follow-up scans according to the RECIST criteria.

Among 67 PROS patients screened from 2008 to 2021, 30 met the inclusion criteria, including 43.3% female patients. The median age at first MRI was 19 years (interquartile range, 5 to 34) and the median interval between the two scans was 75.7 months (range, 2.1 to 160.3 months). Recurrent localizations included the face (n = 6; 20%), ear, nose and throat region (n = 3; 10%), upper limbs (n = 5; 16.7%), thorax (n = 3; 10%), abdomen (n = 4; 13.3%), pelvis (n = 5; 16.7%), and lower limbs (n = 10; 33.3%), with some patients presenting multisite involvement.

During the observation period, 86.6% (n = 26) of patients exhibited an increase in target lesion volume, with a median progression of 37.8% (range, 2.6 to 233.0%) and a mean progression of 52% (standard error of the mean, 7.2%), reflecting a right-skewed distribution driven by a subset of rapidly enlarging lesions. In conclusion, this study provides the first radiological description of the natural history of PROS, demonstrating that tissue malformations most often enlarge over time, with sustained progression persisting into adulthood.

## Introduction

*PIK3CA*-related overgrowth spectrum (PROS) encompasses a heterogeneous group of disorders caused by postzygotic activating variants in the *PIK3CA* gene, leading to mosaic hyperactivation of the phosphoinositide 3-kinase (PI3K)/ (AKT)/mammalian target of rapamycin (mTOR) signaling pathway^1^. These conditions include former entities such as CLOVES syndrome, Klippel-Trenaunay syndrome, and megalencephaly-capillary malformation (MCAP) syndrome, among others^1^. Despite clinical heterogeneity, these disorders are all driven by somatic gain-of-function variants in *PIK3CA*. The resulting pathway hyperactivation drives abnormal cell proliferation, angiogenesis, adipogenesis, and tissue hypertrophy in affected regions. The clinical presentations of PROS are remarkably variable and depend largely on the embryogenic stage at which the mutational event occurs, and on how the mutant cells are distributed across tissues^1^. Early embryonic mutations can cause widespread, multisystemic phenotypes, whereas later events tend to produce localized or segmental overgrowth^1,3^. Commonly affected tissues include vascular structures (capillary, venous, and lymphatic), adipose tissue, skeletal muscle, and bone, with occasional involvement of internal organs. In addition to morphological changes, patients frequently suffer from chronic pain, recurrent inflammation, bleeding, oozing, thrombosis, and reduced mobility.^3^ The psychosocial and esthetic impact of these manifestations further contributes to disease burden.

Despite major advances in both PROS pathophysiology and genetic diagnosis, the natural history of the disorder remains poorly defined. Existing knowledge is mainly derived from case reports, small series, or cross-sectional analyses limited to specific phenotypes^4–6^. While these have provided valuable descriptive insights, they offer little information on longitudinal disease progression in untreated individuals. Consequently, clinicians lack robust prognostic data to guide discussions about expected growth trajectories or functional outcomes over time.

The absence of longitudinal data has important clinical consequences. It hinders accurate prognostic counseling for patients and families, and questions about whether lesions will stabilize, regress, or continue to expand often go unanswered. It also complicates therapeutic decision-making. The recent development of targeted therapies, including mTOR^7^ and PI3K inhibitors^8–14^, has transformed PROS management; yet evaluating treatment efficacy requires a clear understanding of spontaneous disease progression. Without natural history data, distinguishing true therapeutic benefit from spontaneous variability remains challenging.

Another limitation lies in the lack of standardized radiological assessments. While magnetic resonance imaging (MRI) is the preferred imaging modality for PROS-related malformations, quantitative longitudinal data on disease progression remain limited. Most available studies rely on qualitative or subjective assessment. Applying standardized oncologic metrics, such as RECIST^15^ (Response Evaluation Criteria in Solid Tumors) to nonmalignant overgrowth disorders could introduce much-needed objectivity and reproducibility.

A further gap concerns the adult population. Although PROS originates in early development, its evolution in adulthood is poorly characterized. Whether lesion growth stabilizes or progresses after puberty remains unclear.

Given these gaps, longitudinal studies of untreated patients with molecularly confirmed PROS are urgently needed. Establishing such natural history data would help quantify progression rates, identify tissues most prone to expansion, and clarify the relationship between imaging and clinical outcomes. It would also provide a necessary baseline for clinical trials and assessment of therapeutic efficacy.

In this context, we conducted a retrospective longitudinal study of pediatric and adult patients with genetically confirmed PROS from two tertiary referral centers. By analyzing serial MRI scans obtained over extended intervals and excluding patients having received surgical, radiological, or pharmacological interventions, we aimed to characterize the intrinsic growth dynamics of PROS lesions. To our knowledge, this study provides the first systematic, quantitative description of the natural history of PROS across a broad age spectrum.

## Materials and methods

### Study Design and Participants

PROS patients referred to two French centers were retrospectively studied. Inclusion criteria were the presence of a genetically confirmed *PIK3CA* mutation, availability of two MRIs performed at least 2 months apart, and a baseline target lesion >1 cm in greatest dimension. Patients having received either interventional (radiological or surgical) procedures or targeted therapies between the two MRI scans were excluded.

### Imaging

MRI scans were performed on 1.5-T or 3.0-T systems using T1-weighted and fat suppressed T2-weighted images (T2FS) using either short tau inversion recovery (STIR), fat saturation (Fat-Sat), spectral attenuated inversion recovery (SPAIR) or Dixon method.

### Data Collection

Patient demographic data and genetic testing were collected. The greatest malformation dimension was measured on T2FS sequences as defined by RECIST^15^ (Response Evaluation Criteria in Solid Tumors). Lesion progression was defined as a ≥20% increase compared to baseline.

### Ethics

This retrospective was approved by the ethical committee INSERM-RHU COSY (IRB approval 2021-A02719-32)

### Statistical Analysis

Variables were described as percentages for qualitative variables; mean (m) and standard deviation (SD) for quantitative variables. Relationships between variables were explored using Fisher’s exact test for qualitative variables and Student’s t-test for quantitative variables. Bilateral significance threshold was set at 5% and 95% confidence intervals were calculated. Differences between the experimental groups were evaluated using Mann–Whitney tests when two groups were compared. Statistical analyses were performed using GraphPad Prism software (version 10.2.0).

## Results

### Cohort characteristics

Among 67 screened patients with genetically confirmed PROS, 30 had a measurable target lesion and ≥2 MRI examinations of the lesion performed at least 2 months apart (**Figure 1**). There was 17 males and 13 females. Median age at first MRI was 19.0 years (range, 5 to 72 years), indicating that most patients were imaged in childhood, adolescence, or early adulthood, with a minority of patients presenting or being reevaluated in later adult life (**Table 1**). This broad age range suggests both early recognition of extensive congenital malformations and delayed diagnosis or late progression in some adults.

**Figure 1.** Flow-chart of the study.

**Table 1:**
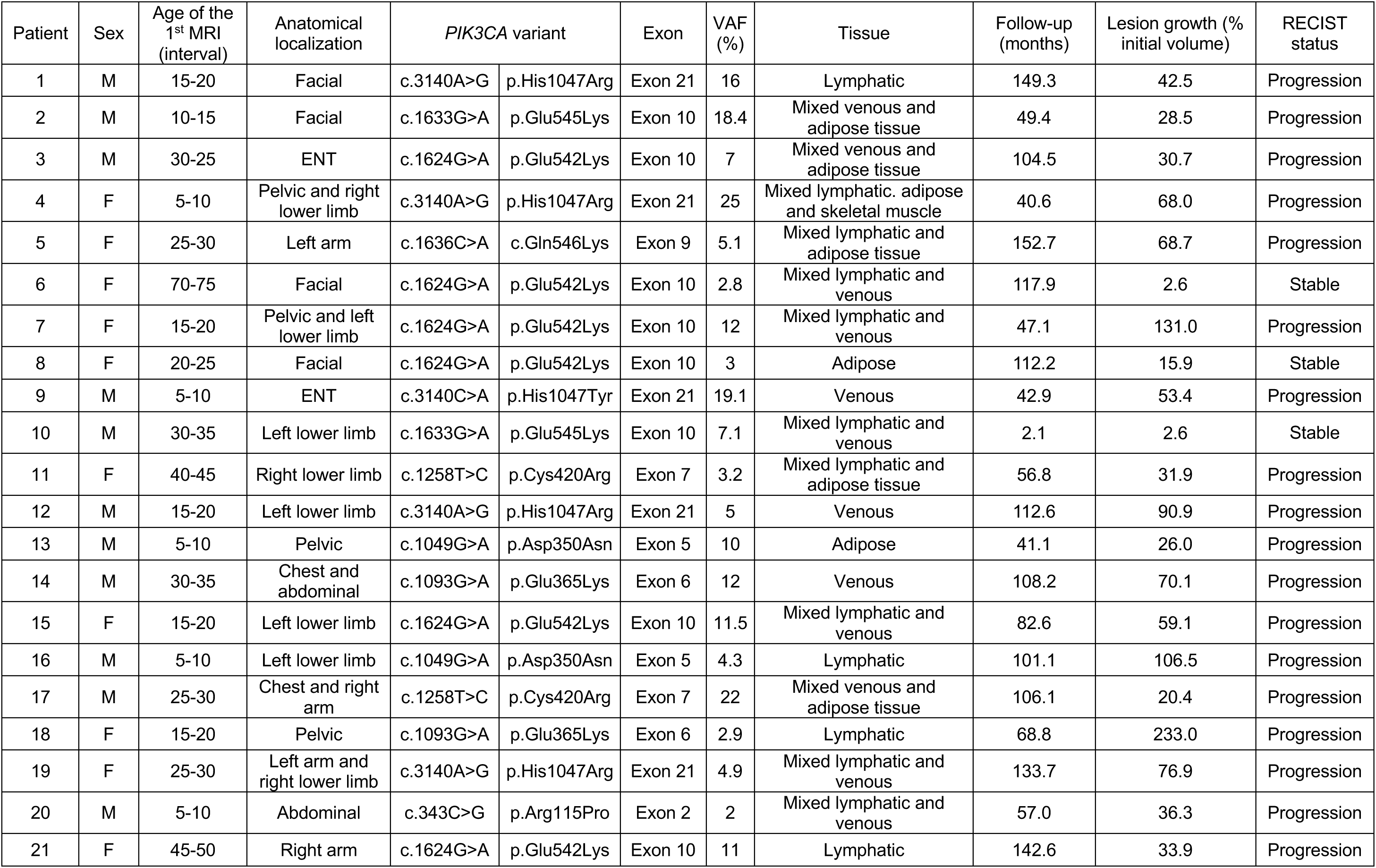
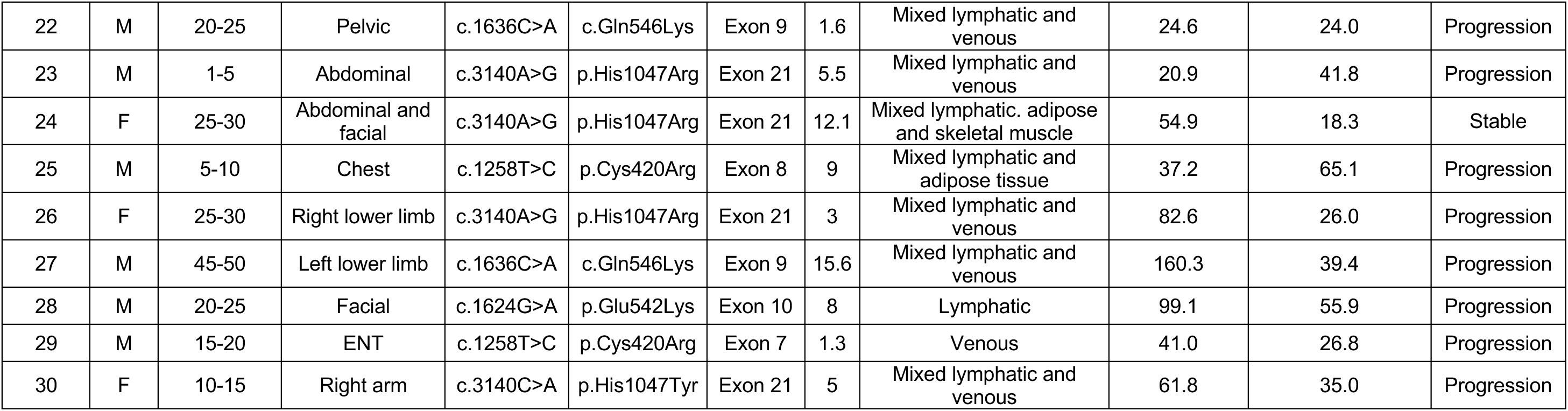
Patients’ characteristics. . MRI: Magnetic Resonance Imaging. VAF: Variant Allele Frequency.

Anatomical localization was heterogeneous, with a predominance of craniofacial and limb involvement, often with segmental distribution. Nine patients (30%) presented with facial (n=6) or ENT (n=3) involvement. Abdominal involvement was present in 4 patients (13.3%), either isolated or as part of combined chest-abdominal disease. Pure pelvic involvement was recorded in 3 patients (10%) and combined pelvic-lower limb (left or right) involvement in 2. Limb lesions were frequent, particularly in the lower extremities: left lower limb involvement occurred in 6 patients (e.g., isolated left lower limb or combined with pelvic disease), right lower limb lesions in at 3 patients, and additional cases had left arm, right arm, or combined arm–lower limb disease. Some patients thus exhibited complex segmental overgrowth patterns encompassing contiguous regions (e.g., pelvis and right lower limb, chest and right arm or chest and abdomen), consistent with mosaic involvement of an embryonic germ layer rather than a single tissue.

Genetically, the most commonly *PIK3CA* affected exons (**Table 1**) were exon 21 and exon 10 (9 patients each), followed by exons 9 and 7 (3 patients each), and exons 6 and 5 (2 patients each), whereas exons 8 and 2 were represented in single patients. 50% of the patients exhibited hotspot *PIK3CA* variants (p.Glu542Lys, n=7; p.His1047Arg, n=6; p.Glu545Lys, n=2). Additional recurrent variants included p.Cys420Arg (n=4), p.Gln546Lys (n=3), p.Asp350Asn (n=2), p.Glu365Lys (n=2), and p.His1047Tyr (n=2), alongside a single p.Arg115Pro variant in exon 2. Variant allele frequencies in the affected tissue ranged from 1.3% to 25.0%, with a median of 7.1%, reflecting the variability of mutant cell burden across patients that likely contributes to the heterogeneity of clinical and radiological phenotypes.

### MRI tissue composition and baseline morphology

Baseline MRIs showed various phenotypes across patients, with a predominance of mixed malformations over single-component lesions (**Table 1**). The most frequent MRI findings were combined venous-lymphatic malformations (n=11, 36.7%), characterized radiologically by serpiginous or multiloculated T2-hyperintense structures and septated spaces consistent with lymphatic channels, interwoven with venous lakes or ectatic veins, sometimes extending along fascial planes and neurovascular bundles (**Figure 2**). Mixed lymphatic and adipose lesions were identified in 3 patients; in these cases, MRI commonly demonstrated disproportionate fatty overgrowth of a segment or region, with embedded fluid-filled cystic spaces or septated areas corresponding to lymphatic malformation within hypertrophic subcutaneous fat. Three additional patients had mixed venous and adipose lesions, combining lobulated fatty overgrowth with dilated venous structures, often leading to contour deformity and asymmetry of the affected limb or trunk region.

**Figure 2.**
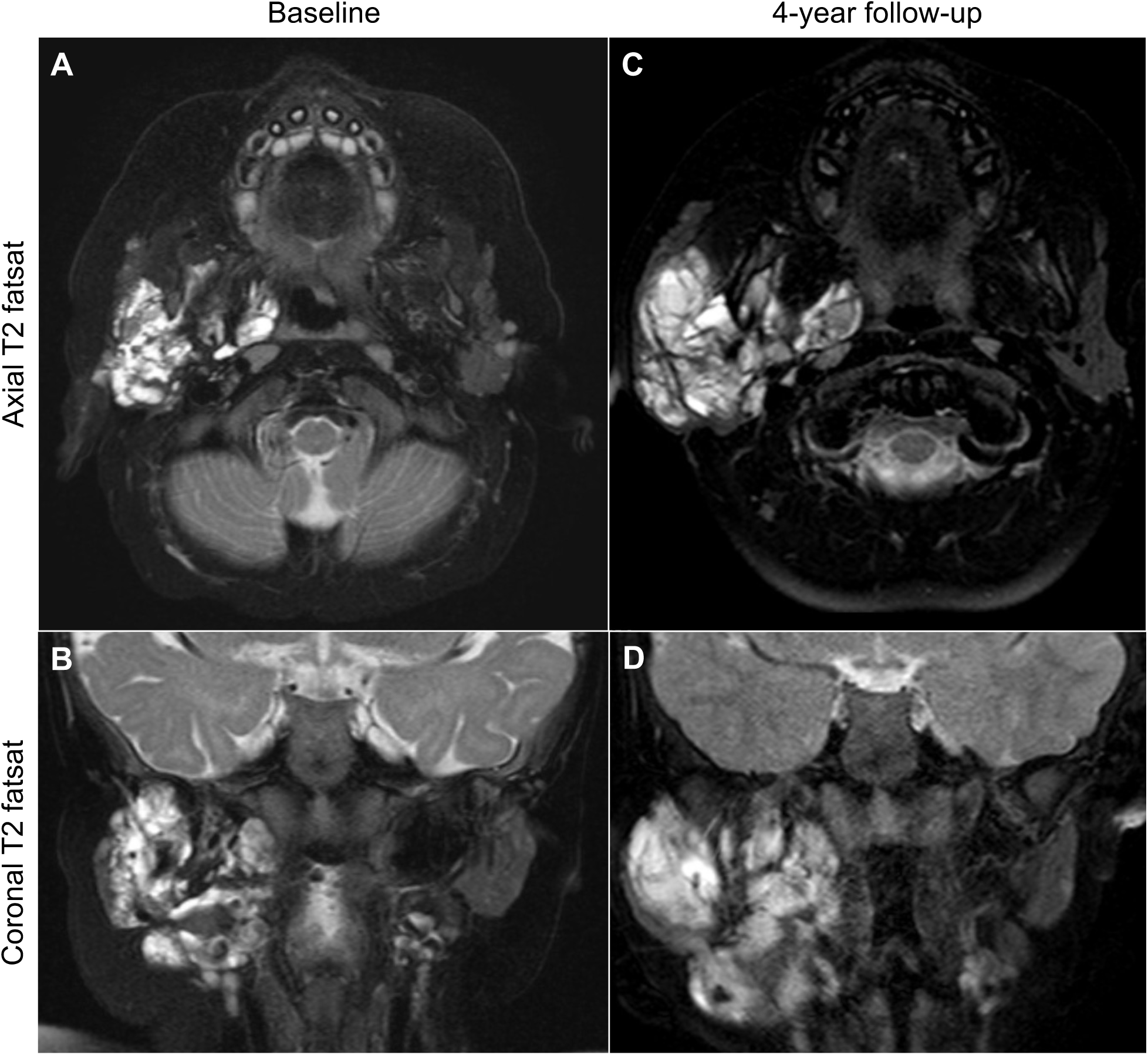
Evolution of a right parotid lymphatic malformation over 49.4 months in patient 2. The lesion increased by 28.5% during follow-up.

Two patients had lesions described as mixed lymphatic, adipose, and skeletal muscle, representing the most complex pattern of tissue infiltration. In these cases, MRI suggested deep extension into or between muscle groups, with replacement or disorganization of normal muscular architecture by high-signal, lobulated tissue on fluid-sensitive sequences and interspersed adipose strands. Purely lymphatic lesions, present in 5 patients, generally manifested as multiloculated, thin-walled cystic structures, predominantly hyperintense on T2-weighted images and hypo- to isointense on T1-weighted images, sometimes with fluid–fluid levels or septal enhancement but without a substantial venous or fatty component. Venous-predominant lesions were identified in 4 patients and were characterized by dilated venous channels or lakes, occasionally with suspected phleboliths, and a tendency to track along known venous pathways rather than forming isolated cystic masses.

The anatomical distribution of these tissue patterns mirrored the clinical heterogeneity of the cohort. Craniofacial and ENT lesions frequently combined lymphatic and venous or venous and adipose components (for example, facial lesions with mixed venous–adipose composition or ENT lesions with venous or mixed venous–adipose appearances), reflecting the complex vascular anatomy and soft-tissue composition of the head and neck. Limb and pelvic lesions often exhibited mixed lymphatic–venous or mixed lymphatic–adipose patterns, especially in segmental overgrowth of the lower limbs where subcutaneous tissue, muscle, and deep fascia are simultaneously involved. Chest and abdominal lesions were similarly variable, ranging from predominantly venous lesions within the chest and abdominal wall to mixed venous–adipose lesions spanning the chest and arm or chest and abdomen. Overall, these baseline MRI features emphasize that in this cohort, *PIK3CA*-related lesions rarely conform to a single classic vascular malformation category, but instead encompass complex malformations with overlapping lymphatic, venous, and fatty components distributed across multiple tissue planes.

### Longitudinal MRI follow-up and volumetric evolution

All 30 patients underwent at least two MRI examinations of the same lesion, enabling quantitative assessment of lesion behavior over time. The median interval between the two MRI examinations was 75.7 months (approximately 6.3 years), with substantial variability across patients (range, 2.1 to 160.3 months). This heterogeneity in imaging follow-up may reflect differences in disease severity or progression rates among patients. Target lesion volume increased by a median of 37.8%, on MRI 2 compared to MRI 1 (**Figures 2 and 3**), with a wide range (2.6% to 233.0%) indicating that progression rates varied substantially across patients.

**Figure 3.**
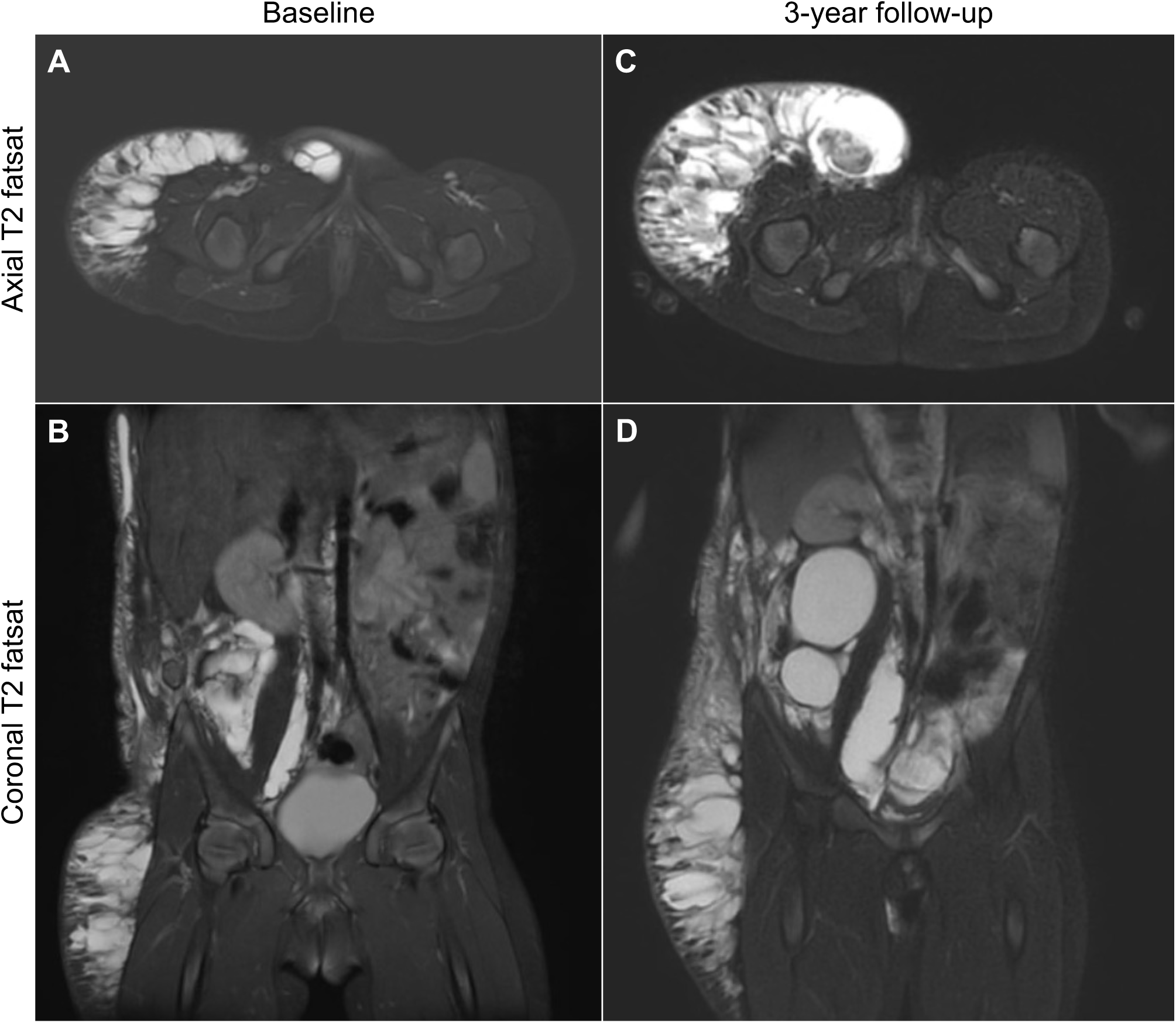
Evolution of a right hypodermic lymphatic malformation over 40.6 months in patient 4. The lesion increased by 68% during follow-up.

To decipher the factors associated with disease progression, we then classified the target lesions as progressive or stable based on the RECIST criteria. Twenty-six patients (86.7%) showed disease progression while only four (13.3%) had stable disease, confirming that PROS are continuously evolving disorders (**Figure 4**). Notably, progression was observed across all malformation types. The most pronounced progression rates were seen in younger patients with lymphatic or mixed lesions, where small baseline volumes may have amplified the relative impact of subsequent growth. In contrast, PROS-related malformations tended to show lower progression rates in older adults.

**Figure 4.**
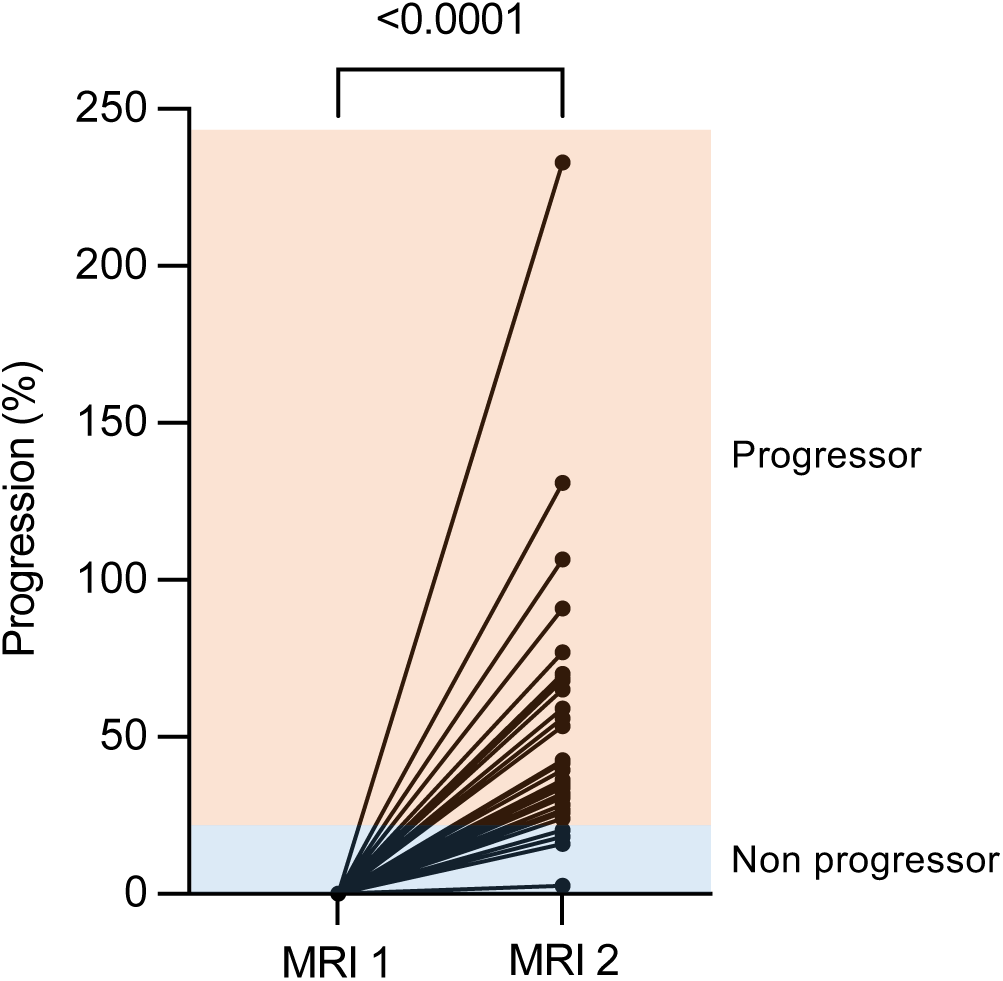
Malformation evolution between 2 MRI.

Malformation burden is not purely volumetric and anatomic context is critical, as lesions in constrained compartments (such as the head and neck, pelvis, or chest) may compromise organ function. However, we observed no clear dichotomy in growth rates between craniofacial, limb, and truncal malformations.

Progression rate also appeared independent of the *PIK3CA* variant involved, as hotspot (involving codons 542, 545 and 1047) and non-hotspot variants both associated with disease progression.

Overall, our data illustrate that PROS-related malformations are structurally complex and display almost systematic, spontaneous progression over time - regardless of tissue involvement and anatomical locations - with a wide variability in growth rates.

## Discussion

This study provides the first longitudinal, imaging-based characterization of the natural history of *PIK3CA*-related overgrowth spectrum (PROS) in untreated patients. By systematically assessing serial MRI scans and applying RECIST criteria to predefined target lesions, we show that PROS lesions most often exhibit intrinsic progression over time. In our cohort, 93.3% of patients demonstrated measurable lesion growth, with great variability in progression rates across patients.

The distribution of volumetric changes was heterogeneous and right-skewed, with a median and a mean increase in lesion volume of approximately 36% and 50%, respectively, over a median follow-up of 6 years. This pattern indicates that while many lesions enlarged moderately, a subset underwent marked expansion over the follow-up period. These findings confirm that PROS-related malformations are continuously progressive conditions rather than stable diseases. However, the presence of two patients with non-progressive lesions highlights inter-individual variability in natural history.

Progression was not uniform across tissue types, but certain patterns emerged. Lesions with a prominent vascular component, including venous, lymphatic, and mixed vascular-adipose malformations, tended to show higher progression rates than predominantly adipose or muscular lesions, although the small sample size limits definitive statistical comparisons. This observation is consistent with the central role of the PI3K pathway in endothelial proliferation and vascular remodeling and may explain the high symptom burden often associated with vascular involvement. Future studies with larger cohorts and formal subgroup analyses will be needed to confirm and refine tissue-specific growth trends.

Similarly, malformation progression was observed regardless of the anatomical localization. However, the clinical consequences of malformation growth likely differ substantially depending on the affected site and surrounding structures. Indeed, modest volumetric increases may have significant functional or esthetic impact in anatomically constrained or visible regions such as the head and neck, pelvis, or hands and feet.

Our observations also inform on the evolution of PROS-related malformations after the end of patients’ growth. By including both pediatric and adult patients, we found that malformations progress beyond childhood and adolescence. While progression rates tended to be higher in the youngest patients - partly due to smaller baseline volumes - our data do not support the concept of overgrowth arrest after puberty. Instead, they suggest that PI3K pathway activation may exert long-lasting biological effects, with a persistent, albeit variable, propensity for lesion expansion throughout life.

From a therapeutic standpoint, defining the natural growth rate and variability of untreated PROS is crucial. Our data provide a quantitative benchmark against which the impact of targeted therapies, such as mTOR or PI3K inhibitors, can be assessed. In a disease that otherwise shows a strong tendency toward enlargement, mild reduction or even stabilization of lesion size may represent a meaningful therapeutic benefit. Our data therefore offer a framework for setting realistic expectations, designing clinical trials, and interpreting treatment responses.

A key strength of this work lies in its rigorous inclusion criteria. Only patients with post-zygotic *PIK3CA* variants were included, which rules out diagnostic uncertainty and reduces phenotypic heterogeneity. By further excluding individuals who underwent surgical, radiological, or pharmacological interventions between MRI assessments, we were able to capture the unmodified natural trajectory of PROS-related malformations over time. This design minimizes confounding effects related to treatment and provides a clearer view of the intrinsic biological behavior of these malformations.

Methodologically, the application of RECIST criteria to nonmalignant conditions represents an important step toward standardized radiological assessment in PROS. Although RECIST was originally developed for solid tumors and may not fully capture the complexity of diffuse or multifocal malformations, it provides a simple and reproducible metric for tracking predefined target lesions over time. Combining RECIST-based measurements with more advanced volumetric or quantitative MRI techniques in future studies could enhance sensitivity to detect subtle changes and better assess disease burden in patients with extensive or multifocal lesions.

The main limitations of this study derive from its retrospective design and relatively small sample size, which are inherent challenges in rare disorders such as PROS. The heterogeneity of lesion types, anatomical sites, and follow-up durations further complicates detailed subgroup analyses and may limit finer genotype-phenotype or tissue-specific patterns. Additionally, while clinical observations suggested that symptom burden often increased alongside radiological progression, systematic collection of standardized clinical and patient-reported outcome measures was not available in all patients. Such considerations will be essential in future prospective works to robustly quantify structure-function relationships.

Despite these limitations, our findings have important implications for clinical practice. They support PROS as progressive mosaic overgrowth syndromes in which lesions most often enlarge over time, including during adulthood. This conceptual shift underscores the need for proactive, longitudinal monitoring and timely consideration of therapeutic strategies, particularly for lesions in functionally critical locations. Establishing individualized surveillance plans and discussing the likelihood of progression with patients and families are key components of comprehensive care.

Future research should aim to integrate imaging biomarkers, molecular profiling, and standardized clinical outcome measures into longitudinal registries and prospective trials. Such efforts will be essential to identify predictors of rapid progression, refine risk stratification, and determine whether early targeted intervention can alter the natural course of PROS and reduce long-term morbidity. Ultimately, a better understanding of the natural history of PROS will provide the foundation for optimizing treatment timing, improving patients’ quality of life, and transforming outcomes.

## Data Availability

All data produced in the present study are available upon reasonable request to the authors

